# Race and Ethnicity Disparities in the Life’s Essential 8 Ever-Pregnant Adults in the United States: The National Health and Nutrition Examination Survey 2011-2020

**DOI:** 10.1101/2024.08.28.24312682

**Authors:** Khadijat Adeleye, Tosin Tomiwa, Yaa Adoma Kwapong, Ellen Boakye, Oluwalonimi Adebowale, Brenda Owusu, Ruth-Alma Turkson-Ocran, Yvonne Commodore-Mensah, Oluwabunmi Ogungbe

## Abstract

**Background:** Cardiometabolic conditions are among the leading causes of maternal mortality in the US. The American Heart Association (AHA) *Life’s Essential 8*^TM^ (LE8) provides an actionable summary measure for assessing cardiovascular health.

**Methods:** We conducted a cross-sectional analysis of National Health and Nutrition Examination Survey (NHANES) data among ever-pregnant adults from 2011 through March 2020. The exposure of interest was race/ethnicity. Primary outcomes included LE8 scores (health outcome and lifestyle metrics). We fitted survey-weighted linear and multinomial logistic regression models, examining racial and ethnic disparities by LE8 scores and each metric separately, adjusting for confounders.

**Results:** Among 2,208 ever-pregnant adults, the mean age was 52.0 ± 19.64 years. Non-Hispanic (NH) Black adults had lower mean LE8 scores (57.20 95%CI: 55.96, 58.44) compared to NH White (62.85 95% CI: 61.39, 64.30), Mexican/Hispanic (62.26, 95%CI 60.86, 63.66), and NH Asian adults (65.83 95% CI: 63.47, 68.19). After adjusting for confounders, NH Black adults had significantly lower overall LE8 scores than NH White adults (β = -0.09, 95% CI: -0.12, -0.06), with lower scores for blood pressure (β = -0.25, 95% CI: -0.32, -0.18) and BMI (β = -0.21, 95% CI: -0.30, -0.11).

Mexican/Hispanic adults were less likely to be in the high LE8 score category (Quartile 3) compared to NH White adults (PRR: 1.32, 95% CI: 0.92 1.91) and had lower physical activity scores (β = -0.38, 95% CI: -0.55, -0.21). NH Asian adults had lower scores for physical activity (β = -0.76, 95% CI: -1.10, -0.4) but higher scores for BMI (β = 0.31, 95% CI: 0.23, 0.40).

**Conclusion:** NH Black, Hispanic ever-pregnant adults had a higher prevalence of adverse cardiometabolic outcomes. Focused interventions are needed to address these disparities and improve maternal cardiometabolic health, per AHA’s LE8 goals.

**Clinical Perspective:** *What Is New?:*

- The Life’s Essential 8 (LE8) score provides a comprehensive and actionable tool for assessing cardiovascular health in ever-pregnant adults, offering clinicians a standardized method to identify and stratify cardiovascular risk.
- Significant racial and ethnic disparities exist in LE8 scores among ever- pregnant adults, with NH Black women consistently showing lower scores across various components, indicating a higher burden of cardiovascular risk factors.
- Higher education levels and socioeconomic status are strongly associated with better LE8 scores, highlighting the importance of addressing social determinants of health in cardiovascular risk management.

*What Are the Clinical Implications?:*

- LE8 score can be used alongside existing risk assessment tools to better identify women at high risk for cardiometabolic complications during pregnancy. This allows for earlier intervention and potentially improved maternal health outcomes.
- For women identified with lower LE8 scores, early intervention becomes crucial.
- Preconception care programs can help optimize their cardiovascular health before pregnancy by promoting healthy diets, physical activity, and weight management.
- The link between lower LE8 scores and lower socioeconomic status underscores the importance of addressing social determinants of health.

## INTRODUCTION

Cardiometabolic conditions are among the leading causes of maternal mortality in the US, accounting for 33% of pregnancy-related deaths,^1^ with (N) Black women facing a 2.9 times higher mortality rate than NH White women.^2^ Examining the American Heart Association’s (AHA) *Life’s Essential 8* (LE8) profile in adults who have ever been pregnant (“ever-pregnant adults”) may be crucial for understanding long-term cardiovascular risk. Women with pregnancy-related cardiometabolic complications face a 2- to 3-fold increased risk of cardiovascular events within 5 to 10 years postpartum,^3,4,5,6^ underscoring the need for targeted interventions to improve cardiovascular health among women of reproductive age.

The AHA’s LE8 metric provides a comprehensive assessment framework for promoting ideal cardiovascular health.^7^ While the importance of these factors in reducing cardiovascular disease (CVD) burden is well-established,^7–9^ there remains a significant gap in our understanding of how this metric manifests among adults with a pregnancy history. This gap is particularly concerning given the known increased cardiovascular risks associated with pregnancy complications^10–12^. Exploring the intersection of race/ethnicity, pregnancy history, and CVD metrics is crucial to understanding the complex interplay of these factors and identifying high-risk populations. Such insights are essential for developing targeted interventions and policies to address disparities and improve long-term cardiovascular health outcomes for women across diverse racial and ethnic groups.

In this study, we utilized data from the National Health and Nutrition Examination Survey (NHANES)^13^ from 2011 to March 2020 to examine the CVH metrics of ever- pregnant adults using the AHA LE8 metrics by race and ethnicity. We evaluated the age-adjusted mean LE8 scores among ever-pregnant adults. We also examined additional determinants that contribute to cardiovascular health disparities among various racial and ethnic populations who have been pregnant in the past.

## METHODS

A detailed description of the survey is available at: http://www.cdc.gov/nchs/nhanes.htm http://www.cdc.gov/nchs/nhanes.htm.

### Participants

Among the 45,462 participants in NHANES from 2011 to 2020, 17,800 younger than 18 were excluded. Participants who were never pregnant and those with missing data on relevant covariates (n=23,542) were excluded; among those who were remaining, an additional 1912 were excluded for incomplete LE8 data. This study included a sample of 2,208 ever-pregnant adults (**Figure S1**). Adults (>18 years old) were considered to have a history of pregnancy if they reported “Yes” to the question of ever being pregnant.

### Exposures

Main exposures, race, and ethnicity were categorized into five groups: non-Hispanic (NH) White, NH Black, Mexican American/Other Hispanic, NH Asian, and other races/ethnicities. The ‘other’ races/ethnicities group was racially/ethnically diverse, including multi-racial/ethnic individuals and those not self-identifying as any of the above-listed races.^8,15^

### Outcomes

The outcome of interest, AHA LE8 quantifies cardiovascular health, which includes 4 health behaviors (diet, physical activity, nicotine exposure, and sleep health) and 4 health factors (body mass index, blood lipids, blood glucose, and blood pressure), to significantly enhance guidance on improving cardiovascular health in the general population. The LE8 score is the mean value of the 8 components. LE8 components are scored on a point system and categorized into ideal, intermediate, and low. A detailed calculation of scores for each metric of LE8 is in **Table S1.**

We rated each of the 8 LE8 indicators on a scale from 0 to 100 points and calculated their unweighted average to obtain the total LE8 score. The overall LE8 score was divided into 4 quartiles: Quartile 1(LE8 score: 29.3-53.0), Quartile 2 (LE8 score:53- 56.88), Quartile 3 (LE8 score: 56.88 – 72.5) and Quartile 4 (LE8 score: ≥72.5). The AHA recommends categorizing LE8 scores >80 as high cardiovascular health, 50 to 79 as moderate cardiovascular health, and <50 as low cardiovascular health.^7^

### Covariates

Demographic variables included age, sex, income, and health insurance. Variables examined as dichotomous include marital status (currently married/not married, including never married, divorced, widowed, or separated), employment status (employed/unemployed), and insurance status (insured/uninsured).

We stratified age into adjusted categories with 10-year intervals: 20-29 years, 30-39 years, 40-49 years, 50-59 years, 60-69 years, and 70+ years. Educational levels were categorized as ≤high school, some college, and ≥ bachelor’s degree. The poverty-income ratio (PIR) was calculated as family income divided by the federal poverty level; a PIR of 1 indicates a family income at the federal poverty level. PIR was categorized as <1.0, 1–1.99, where ≥2. A PIR of <1 means that the individual income is below the federal poverty level. Between 1 and 1.99 indicates the income is between 100 and 199% of the poverty level, and ≥2 means that the income is more than 200% of the federal poverty level.

### Statistical Analysis

We examined sociodemographic characteristics using descriptive statistics, including means, standard deviations, and percentages. We computed age-standardized mean scores, standard errors (SE), and 95% confidence intervals for overall LE8 and each LE8 component. Age-standardized mean estimates were calculated using the direct standardization method, with the 2000 US Census population as the standard population. To assess racial and ethnic differences in the cardiometabolic profile of ever-pregnant adults, we modeled LE8 scores continuously, using survey-weighted generalized linear with Gaussian distribution, adjusting for confounders (age, education, income, insurance, and employment). We calculated both unadjusted and adjusted mean differences to assess the impact of confounding variables.

Afterward, we used survey-weighted multinomial logistic regression to assess the association between overall LE8 score quartiles (categorical outcome) and race/ethnicity, education, income, employment, insurance, and marital status. The multinomial logistic regression used the lowest LE8 score quartile as the reference category. We calculated prevalence rate ratios (PRRs) with 95% confidence intervals for each LE8 component and overall LE8 score quartiles. We performed a similar analysis with LE8 score categories of ideal, intermediate, and low, with the ideal category as the reference group. All analyses were performed using STATA 18.^16^ Statistical significance was defined as 2-sided α <0.05.

We conducted a sensitivity analysis excluding participants of ‘other’ races to assess the robustness of our findings. Additionally, we examined the association between LE8 scores and age using categorical age groups to capture non-linear relationships.

## RESULTS

### Sample Characteristics

After we applied survey weights, the sample was 2,208 ever-pregnant adults, representing 13807465 ever-pregnant adults in the US population. The mean age was 52.0 (±19.64 years). **Table 1** presents weighted demographic characteristics across racial and ethnic groups; unweighted distributions are in **Table S2**.

**Table 1:** Weighted Sample Characteristics of Ever-Pregnant Adults by Race and Ethnicity, NHANES 2011-2020.

| Variable n (%) | Non-Hispanic<br>White | Mexican<br>American/Other<br>Hispanic | Non-Hispanic<br>Black | Non-Hispanic<br>Asian | Other Races | Total |
| --- | --- | --- | --- | --- | --- | --- |
| N | 9220359 (66.78) | 1957638 (14.18) | 1606445 (11.64) | 503989 (3.65) | 519034 (3.76) | 13807465(100) |
| Age, yr. (mean, SD) | 54.43(14.70) | 45.42(22.64) | 48.03 (30.05) | 49.79(25.10) | 48.12 (18.96) | 52.00(±19.64) |
| **Age Categories -10yr intervals** |  |  |  |  |  |  |
| 20-29 years | 553903 (6.01) | 322357 (16.47) | 244339 (15.21) | 27000 (5.36) | 54684 (10.54) | 1202282 (8.71) |
| 30-39 years | 1459680 (15.83) | 448932 (22.93) | 306458 (19.08) | 113242 (22.47) | 112239 (21.63) | 2440551 (17.68) |
| 40-49 years | 1585096 (17.19) | 439461 (22.45) | 305844 (19.04) | 112326 (22.29) | 132522 (25.53) | 2575249 (18.65) |
| 50-59 years | 1873487 (20.32) | 388202 (19.83) | 336838 (20.97) | 129755 (25.75) | 73436 (14.15) | 2801717 (20.29) |
| 60-69 years | 1947527 (21.12) | 203489 (10.40) | 245110 (15.26) | 73517 (14.59) | 102423 (19.73) | 2572066 (18.63) |
| 70+ years | 1800667 (19.53) | 155198 (7.93) | 167856 (10.45) | 48150 (9.55) | 43731 (8.43) | 2215602 (16.05) |
| **Education** |  |  |  |  |  |  |
| High School | 2918925 (31.66) | 1059393 (54.12) | 663390 (41.30) | 103924 (20.62) | 225166 (43.38) | 4970798 (36.00) |
| Some College | 3284276 (35.62) | 576407 (29.444) | 573704 (35.713) | 103828 (20.601) | 215705 (41.559) | 4753921 (34.43) |
| College graduate<br>and above | 3017158 (32.72) | 321838 (16.44) | 369351 (22.99) | 296237 (58.78) | 78163 (15.06) | 4082747 (29.57) |
| **Poverty-Income Ratio** |  |  |  |  |  |  |
| PIR<1 | 790231 (8.57) | 502212 (25.65) | 419922 (26.14) | 21199 (4.21) | 108333 (20.87) | 1841898 (13.34) |
| PIR 1-1.99 | 1526466 (16.56) | 591565 (30.22) | 454917 (28.32) | 65480 (12.99) | 143507 (27.65) | 2781934 (20.15) |
| PIR ≥2 | 6903662 (74.87) | 863862 (44.13) | 731606 (45.54) | 417310 (82.80) | 267195 (51.48) | 9183635 (66.51) |
| **Health Insurance Status** |  |  |  |  |  |  |
| No Health Insurance | 528656 (5.73) | 461939 (23.60) | 217841 (13.56) | 33313 (6.61) | 36324 (6.99) | 1278073 (9.26) |
| Have Health<br>Insurance | 8691704 (94.27) | 1495699 (76.40) | 1388604 (86.44) | 470677 (93.39) | 482710 (93.00) | 12529394 (90.74) |
| **Employment Status** |  |  |  |  |  |  |
| Unemployed | 4455529 (48.32) | 887222 (45.32) | 672618 (41.87) | 204773 (40.63) | 221189 (42.62) | 6441331 (46.65) |
| Employed | 4764830 (51.68) | 1070416 (54.68) | 933828 (58.13) | 299216 (59.37) | 297845 (57.39) | 7366136 (53.35) |
| Values are presented as weighted percentages to represent the US population. P-values were <0.001 for all variables except for employment status, for which P-values were 0.24. P-value based on chi-square tests for categorical variables and analysis of variance for continuous variables. |  |  |  |  |  |  |

### Age-Standardized Mean LE8 Scores Among US Ever-Pregnant Adults

There were significant differences in mean LE8 scores among racial and ethnic groups (**Table 2** and **Figure 1**). The overall age-standardized mean of LE8 component scores, including the health outcomes and health behaviors by race and ethnicity, are illustrated in **Figures 2 and 3**, respectively. NH Asian ever-pregnant adults had the highest mean overall LE8 score (65.83,95% CI: 63.47 - 68.19), followed by NH White (62.85,95% CI: 61.39 – 64.30), and Mexican/Hispanic ever-pregnant adults (62.26,95% CI: 60.86 – 63.66) NH Black ever-pregnant adults had the lowest mean score (57.20,95% CI: 55.96 – 58.44). In the sample, NH White ever-pregnant adults had the highest mean score for blood glucose (89.70, 95% CI: 87.59-91.80), while NH Black ever-pregnant adults had the lowest (79.61, 95% CI: 78.86-81.36). For blood pressure, NH White (74.86, 95% CI: 71.89-77.84) and Mexican American/Other Hispanic (74.12, 95% CI: 71.40-76.84) ever-pregnant adults had the highest mean scores, while NH Black individuals had the lowest (58.42, 95% CI: 54.68-62.16).

**Figure 1:**
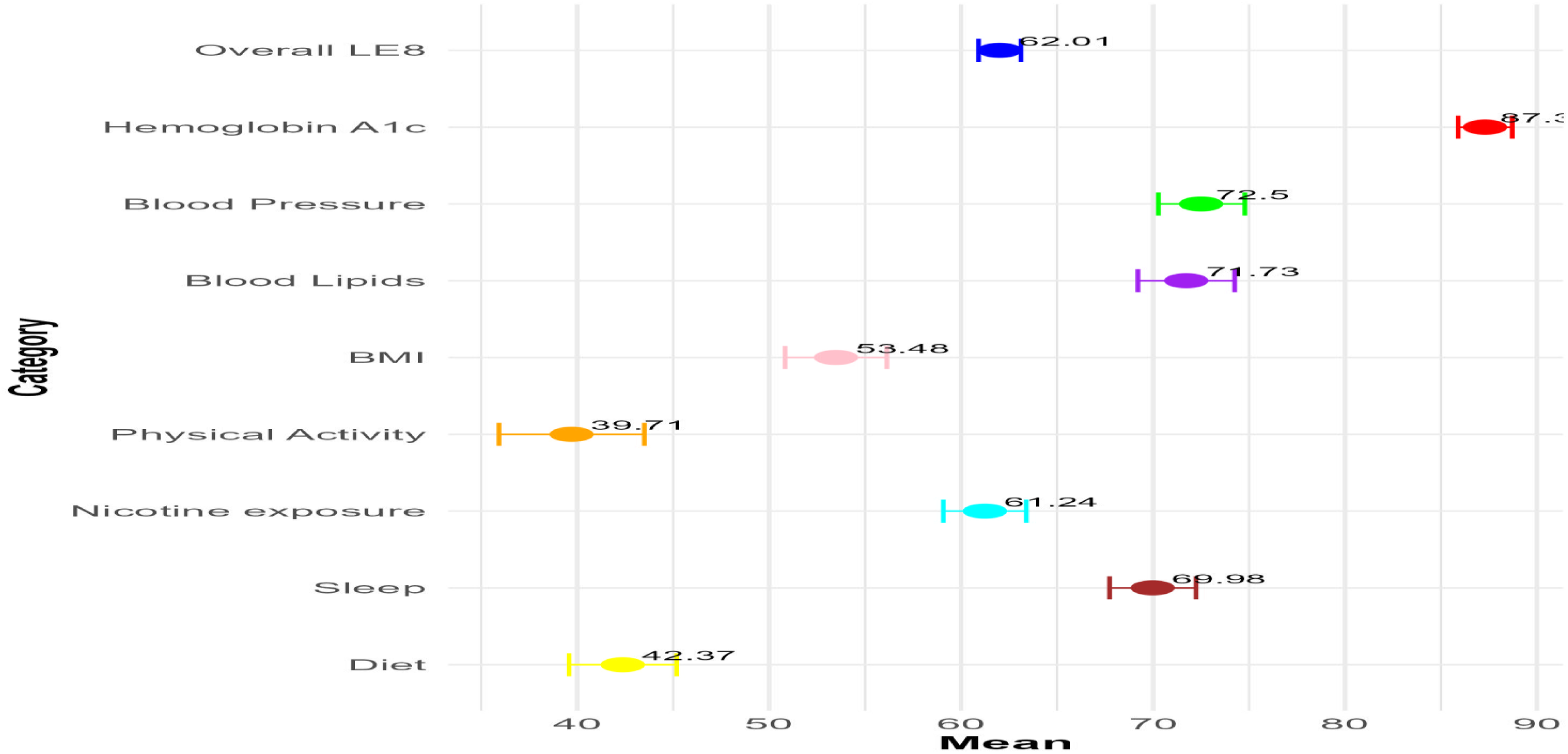
Age-Standardized Mean and 95% Confidence Intervals of Life’s Essential 8 Scores by Race/Ethnicity, NHANES 2011-2020.

**Figure 2:**
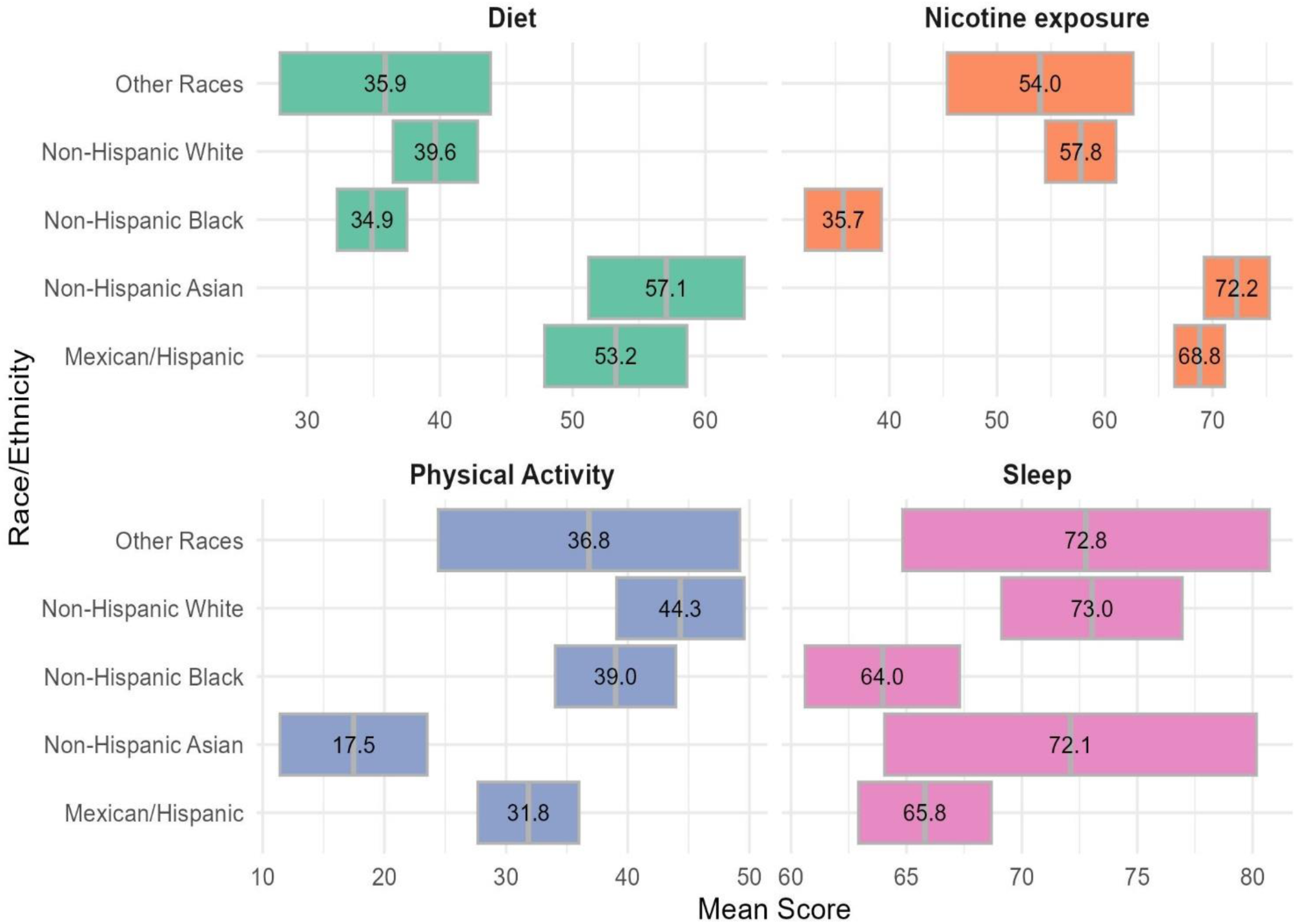
Age-Standardized Mean and 95% Confidence Intervals of Life’s Essential 8 Health Behaviors Scores by Race/Ethnicity, NHANES 2011-2020.

**Figure 3:**
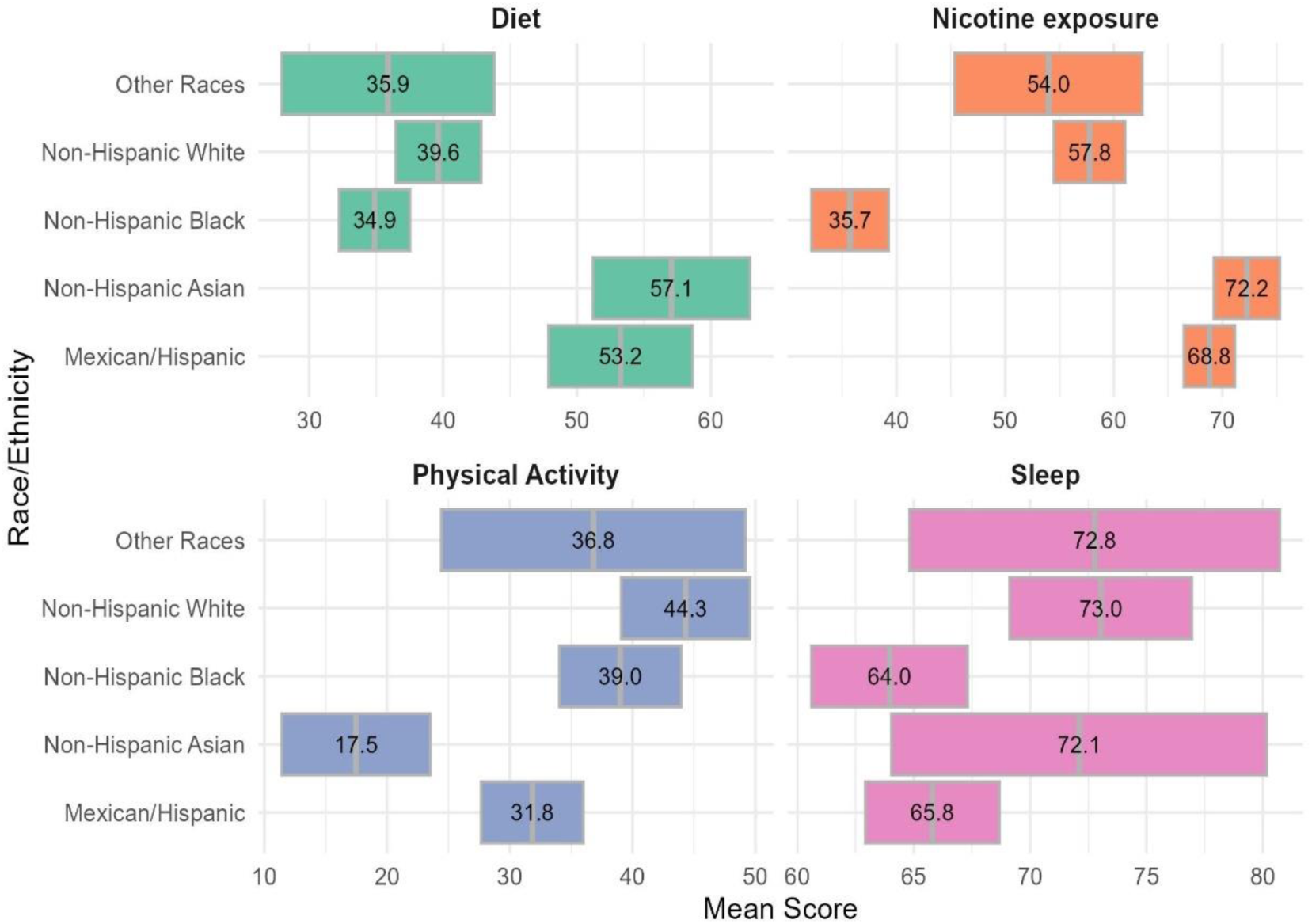
Age-Standardized Mean and 95% Confidence Intervals for Life’s Essential 8 Health Factors Scores by Race/Ethnicity, NHANES 2011-2020.

**Table 2:** Age-Standardized Mean Essential 8 scores Among Ever-Pregnant Adults by Race/Ethnicity, NHANES 2011-2020.

| **AHA LE8 Categories** | **Total** | **Non-Hispanic White** | **Mexican/Hispanic** | **Non-Hispanic Black** | **Non-Hispanic Asian** | **Other Races** |
| --- | --- | --- | --- | --- | --- | --- |
| **Overall LE8** | 62.01<br>(60.91 - 63.12) | 62.85<br>(61.39 - 64.30) | 62.26<br>(60.86 - 63.66) | 57.20<br>(55.96 - 58.44) | 65.83<br>(63.47 - 68.19) | 56.61<br>(53.06 - 60.15) |
| **Hemoglobin A1c** | 87.30<br>(85.89 - 88.71) | 89.70<br>(87.59 - 91.80) | 83.39<br>(81.68 - 85.09) | 79.61<br>(77.86 - 81.36) | 86.17<br>(82.92 - 89.41) | 82.92<br>(78.46 - 87.38) |
| **Blood Pressure** | 72.5<br>(70.27 - 74.78) | 74.86<br>(71.89 - 77.84) | 74.12<br>(71.40 - 76.84) | 58.42<br>(54.68 - 62.16) | 70.59<br>(63.57 - 77.61) | 68.91<br>(57.61 - 80.21) |
| **Blood Lipids** | 71.73<br>(69.21 - 74.25) | 72.13<br>(68.29 - 75.97) | 69.29<br>(66.05 - 72.54) | 77.07<br>(73.76 - 80.38) | 71.65<br>(65.35 - 77.94) | 60.12<br>(48.47 - 71.77) |
| **Physical Activity** | 39.71<br>(35.93 - 43.50) | 44.32<br>(39.07 - 49.57) | 31.84<br>(27.67 - 35.99) | 39.00<br>(34.04 - 43.96) | 17.47<br>(11.42 - 23.52) | 36.81<br>(24.42 - 49.20) |
| **BMI** | 53.48<br>(50.83 - 56.13) | 54.34<br>(50.76 - 57.93) | 52.72<br>(49.58 - 55.87) | 44.01<br>(39.88 - 48.15) | 81.11<br>(75.84 - 86.37) | 46.42<br>(43.35 - 49.49) |
| **Nicotine exposure** | 61.24<br>(59.08 - 63.40) | 57.77<br>(54.48 - 61.05) | 68.81<br>(66.46 - 71.15) | 35.72<br>(32.16 - 39.28) | 72.25<br>(69.22 - 75.28) | 53.99<br>(45.35 - 62.63) |
| **Sleep** | 69.98<br>(67.73 - 72.24) | 73.04<br>(69.12 - 76.95) | 65.80<br>(62.92 - 68.68) | 63.96<br>(60.60 - 67.31) | 72.11<br>(64.05 - 80.17) | 72.78<br>(64.83 - 80.73) |
| **Diet** | 42.37<br>(39.57 - 45.18) | 39.65<br>(36.46 - 42.84) | 53.25<br>(47.88 - 58.62) | 34.88<br>(32.24 - 37.52) | 57.06<br>(51.19 - 62.93) | 35.88<br>(27.94 - 43.81) |
| Values are presented as mean scores with 95% confidence intervals<br>95% confidence intervals in brackets * p<0.05, ** p<0.01, *** p<0.001 |  |  |  |  |  |  |

NH Black ever-pregnant adults had the highest mean score for blood lipids (77.07, 95% CI: 73.76-80.38), while those categorized under Other Races had the lowest (60.12, 95% CI: 48.47-71.77). NH White ever-pregnant adults had the highest mean score for physical activity (44.32, 95% CI: 39.07-49.57), while NH Asian adults had the lowest (17.47, 95% CI: 11.42-23.52). NH Asian ever-pregnant adults had a high mean score for BMI (81.11, 95% CI: 75.84-86.37), while NH Black ever-pregnant adults had the lowest (44.01, 95% CI: 39.88-48.15). For nicotine exposure, NH Asian (72.25, 95% CI: 69.22-75.28) and Mexican American/Other Hispanic (68.81, 95% CI: 66.46-71.15) ever-pregnant adults had the highest mean scores, while NH Black ever-pregnant adults had the lowest (35.72, 95% CI: 32.16-39.28).

For diet, NH Asian ever-pregnant adults reported the highest mean score (57.06, 95% CI: 51.19-62.93), while NH Black ever-pregnant adults had the lowest (34.88, 95% CI: 32.24-37.52). NH White ever-pregnant adults had the highest mean score for sleep (73.04, 95% CI: 69.12 - 76.95), while NH Black individuals had the lowest (63.96, 95% CI: 60.60 - 67.31).

### Racial and Ethnic Differences in Cardiometabolic Profile (LE8 Scores) for Ever- Pregnant Adults

In the unadjusted model of overall LE8 scores, NH Blacks showed significantly lower overall LE8 scores compared to NH White adults (β = -0.09, 95% CI: -0.12, -0.06, **Table 3**), while NH Asian adults had significantly higher scores (β = 0.061, 95% CI: 0.03, 0.09). After adjusting for age, education, income, insurance, and employment, NH Black ever-pregnant adults remain significantly lower than non-Hispanic White adults (β = -0.09, 95% CI: -0.12, -0.06). The Other Races group also had lower scores (β = -0.09, 95% CI: -0.17, -0.01).

**Table 3:**
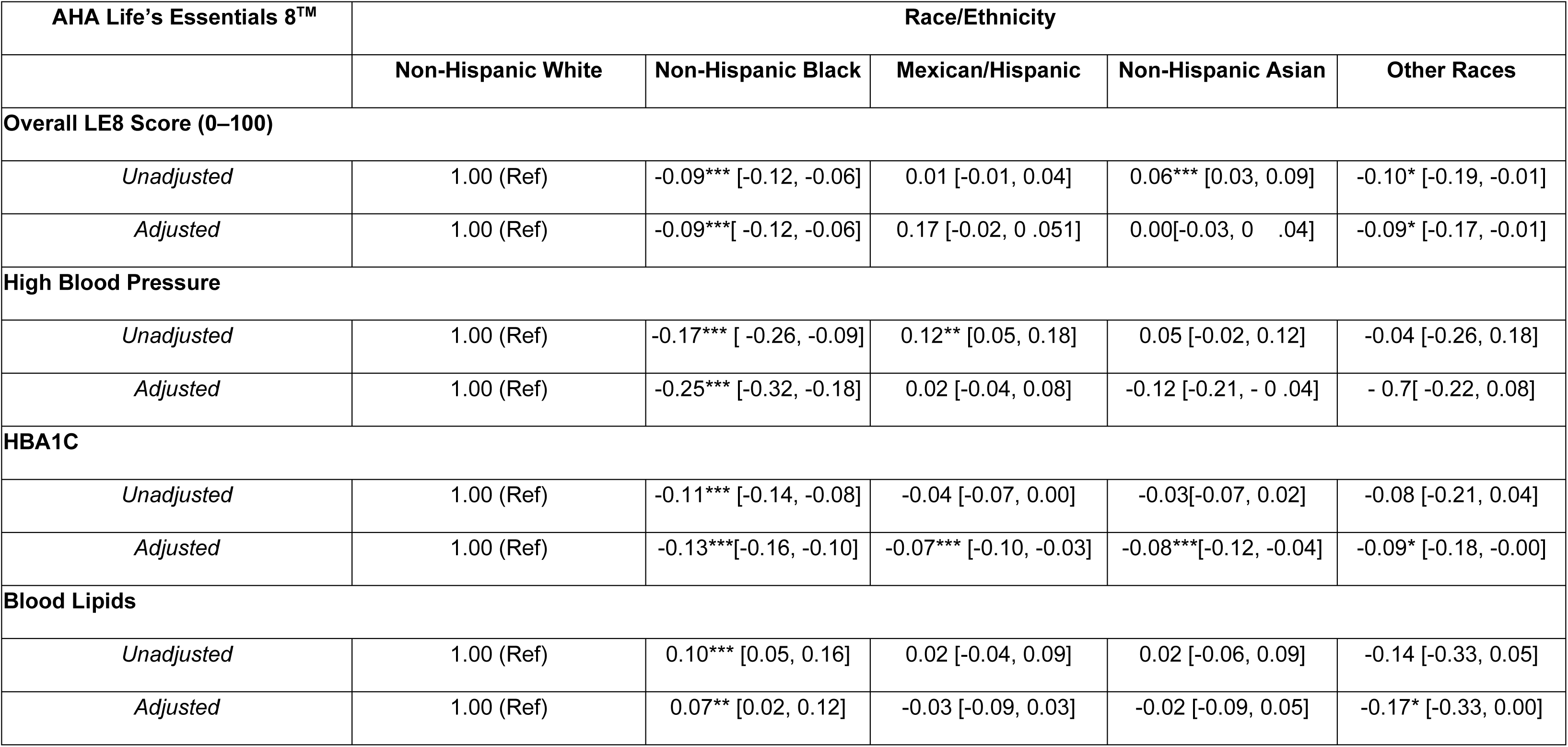
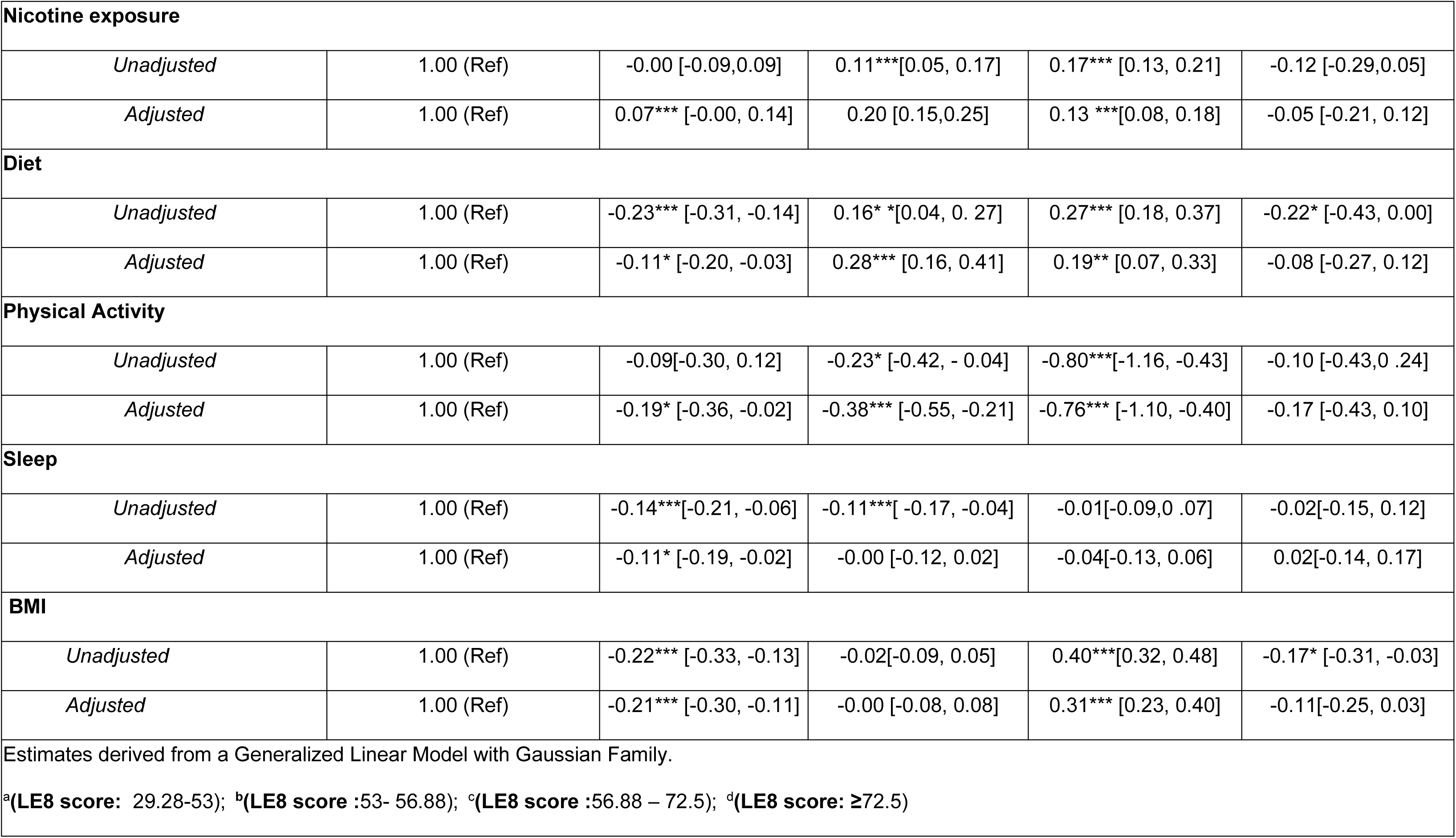
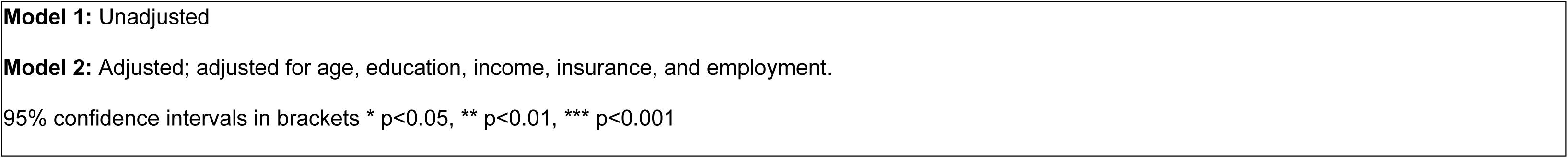
Racial and Ethnic Differences in Life’s Essential 8 Components Among Ever-Pregnant Adults, NHANES 2011-2020.

For blood pressure, NH Black adults had significantly lower blood pressure scores compared to NH White adults (β = -0.25, 95% CI: -0.32, -0.18), indicating a higher likelihood of high blood pressure. All other racial/ethnic groups showed significantly lower hemoglobin A1c (HbA1c) scores compared to NH White adults, with NH Black adults having the largest difference (β = -0.13, 95% CI: -0.16, -0.10), followed by NH Asian (β = -0.08, 95% CI: -0.12, -0.04), Mexican/Hispanic (β = -0.07, 95% CI: -0.10, -0.03), and Other Races (β = -0.009, 95% CI: -0.18, -0.00). NH Black adults had significantly higher blood lipid scores than NH White adults (β = 0.06, 95% CI: 0.01, 0.12).

Mexican/Hispanic adults had significantly higher nicotine exposure scores (indicating less exposure) compared to NH White adults (β = 0.20, 95% CI: 0.15, 0.25). For diet, NH Black adults had significantly lower diet scores compared to non-Hispanic White adults (β = -0.11, 95% CI: -0.20, -0.03), while Mexican/Hispanic (β = 0.28, 95% CI: 0.16, 0.41) and NH Asian adults (β = 0.19, 95% CI: 0.07, 0.33) had significantly higher scores. All other racial/ethnic groups showed significantly lower physical activity scores compared to non-Hispanic White adults, with NH Asian adults showing the largest difference (β = -0.76, 95% CI: -1.10, -0.40), followed by Mexican/Hispanic (β = -0.38, 95% CI: -0.55, -0.21) and NH Black adults (β = -0.19, 95% CI: -0.36, -0.02). NH Black adults had significantly lower sleep scores than NH White adults (β = -0.11, 95% CI: -0.19, -0.02). Finally, for BMI, NH Black adults had significantly lower BMI scores (indicating higher BMI) compared to NH White adults (β = -0.21, 95% CI: -0.30, -0.11), while NH Asian adults had significantly higher scores (indicating lower BMI) (β = 0.31, 95% CI: 0.23, 0.40).

### Racial and Ethnic Differences in LE8 Scores for Ever-Pregnant Adults by Quartiles and Low, Intermediate, and Ideal Categories

Figure 4 shows the relationship between overall LE8 score quartiles and race and ethnicity categories. NH Black adults were less likely to be in the ideal LE8 (Quartile 4) category (PRR: 0.27, 95%CI: 0.16-0.45), Mexican/Hispanic adults were less likely to be in LE8 Quartile 3 (PRR: 1.32, 95%CI: 0.92, 1.91) compared to NH White adults. In comparison, NH Asian adults are likelier to be in LE8 Quartile 3 (PRR: 1.32, 95%CI: 0.92, 1.91).

**Figure 4:**
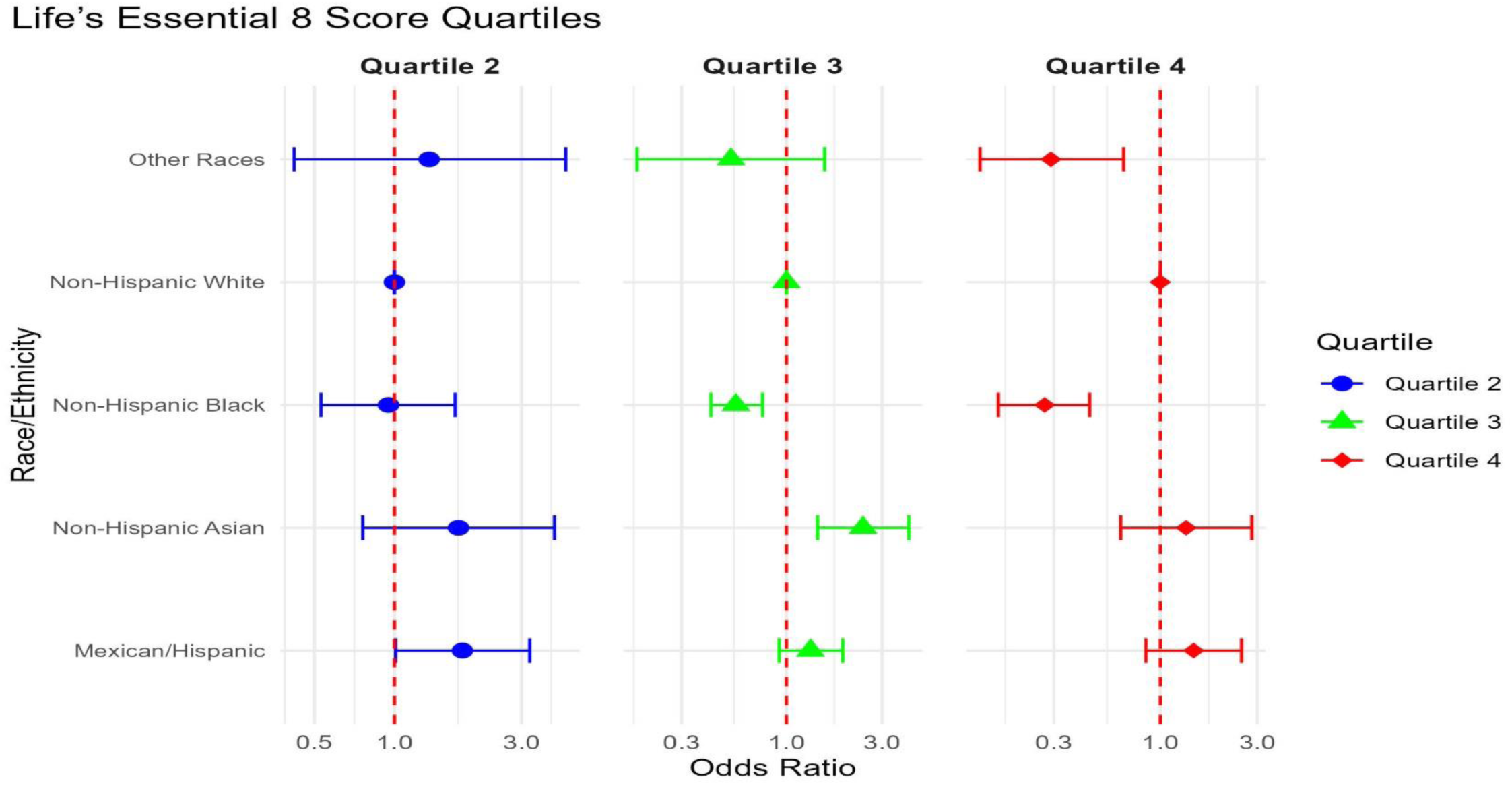
Adjusted Prevalence Rate Ratios of Life’s Essential 8 Score Quartiles by Race and Ethnicity Among Ever- Pregnant Adults (NHANES 2011-2020) Note: The figure displays prevalence rate ratios (PRRs) for being in each Life’s Essential 8 (LE8) score quartile for non-Hispanic Black, non-Hispanic Asian, Mexican/Hispanic, and Other Races ever-pregnant adults, compared to the reference group (non-Hispanic White). Estimates are derived from survey-weighted multinomial logistic regression analyses using NHANES 2011-2020 data. The reference category is Quartile 1 (lowest LE8 scores). PRRs above 1 indicate a higher likelihood, while PRRs below 1 indicate a lower likelihood of being in that quartile than non-Hispanic Whites. Models are adjusted for age, education, poverty-income ratio, health insurance status, and employment. Whiskers represent 95% confidence intervals. LE8 score ranges: Quartile 1 (25-47.5), Quartile 2 (47.5-51.14), Quartile 3 (51.14-67.5), Quartile 4 (≥67.5).

The prevalence rate ratios (PRRs) for the intermediate and low categories of each LE8 component, compared to the ideal category, across different racial/ethnic groups are shown in Figures 5-6 and **Table S3**. For blood pressure, NH Black adults had a significantly higher likelihood of being in the intermediate (PRR: 2.20, 95% CI: 1.67- 2.89) and low (PRR: 3.73, 95% CI: 2.56-5.42) categories compared to NH White adults. Similarly, for blood glucose, NH Black adults showed higher likelihood for both intermediate (PRR: 3.44, 95% CI: 2.85-4.14) and low (PRR: 3.06, 95% CI: 2.05-4.57) categories. For BMI, NH Black adults had a higher likelihood of being in a low category (PRR: 1.90, 95% CI: 1.44-2.52), while NH Asian adults had a significantly lower likelihood (PRR: 0.16, 95% CI: 0.09-0.28) compared to NH White adults. Regarding physical activity, Mexican/Hispanic (PRR: 1.75, 95% CI: 1.26-2.41) and NH Asian (PRR: 2.85, 95% CI: 1.65-4.91) adults were more likely to be in the low category.

**Figure 5:**
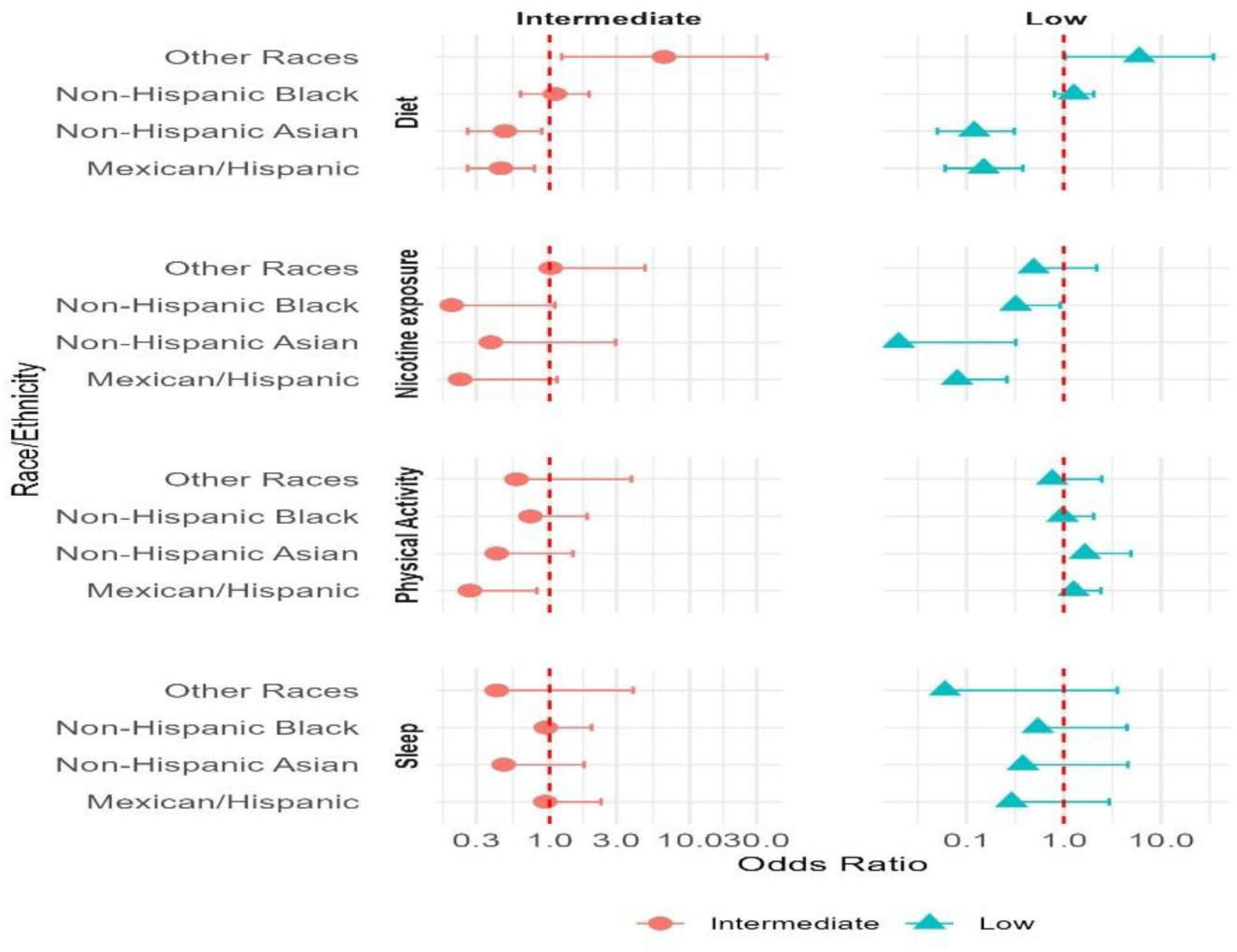
Adjusted Prevalence Rate Ratios of Life’s Essential 8 Health Behavior Components by Race and Ethnicity Among Ever- Pregnant Adults (NHANES 2011-2020) Note: This figure illustrates adjusted prevalence rate ratios (PRRs) with 95% confidence intervals for the health behavior components of the Life’s Essential 8 (LE8) score by race and ethnicity among ever-pregnant adults. The components shown are diet, smoking (nicotine exposure), physical activity (PA), and sleep. Estimates are derived from survey-weighted multinomial logistic regression analyses using NHANES 2011- 2020 data. The reference group is non-Hispanic White adults, and the reference category for each component is the “Ideal” level. PRRs above 1 indicate a higher likelihood, while PRRs below 1 indicate a lower likelihood of being in the “Intermediate” or “Poor” category than non- Hispanic Whites. Models are adjusted for age, education, poverty-income ratio, health insurance status, and employment.

**Figure 6:**
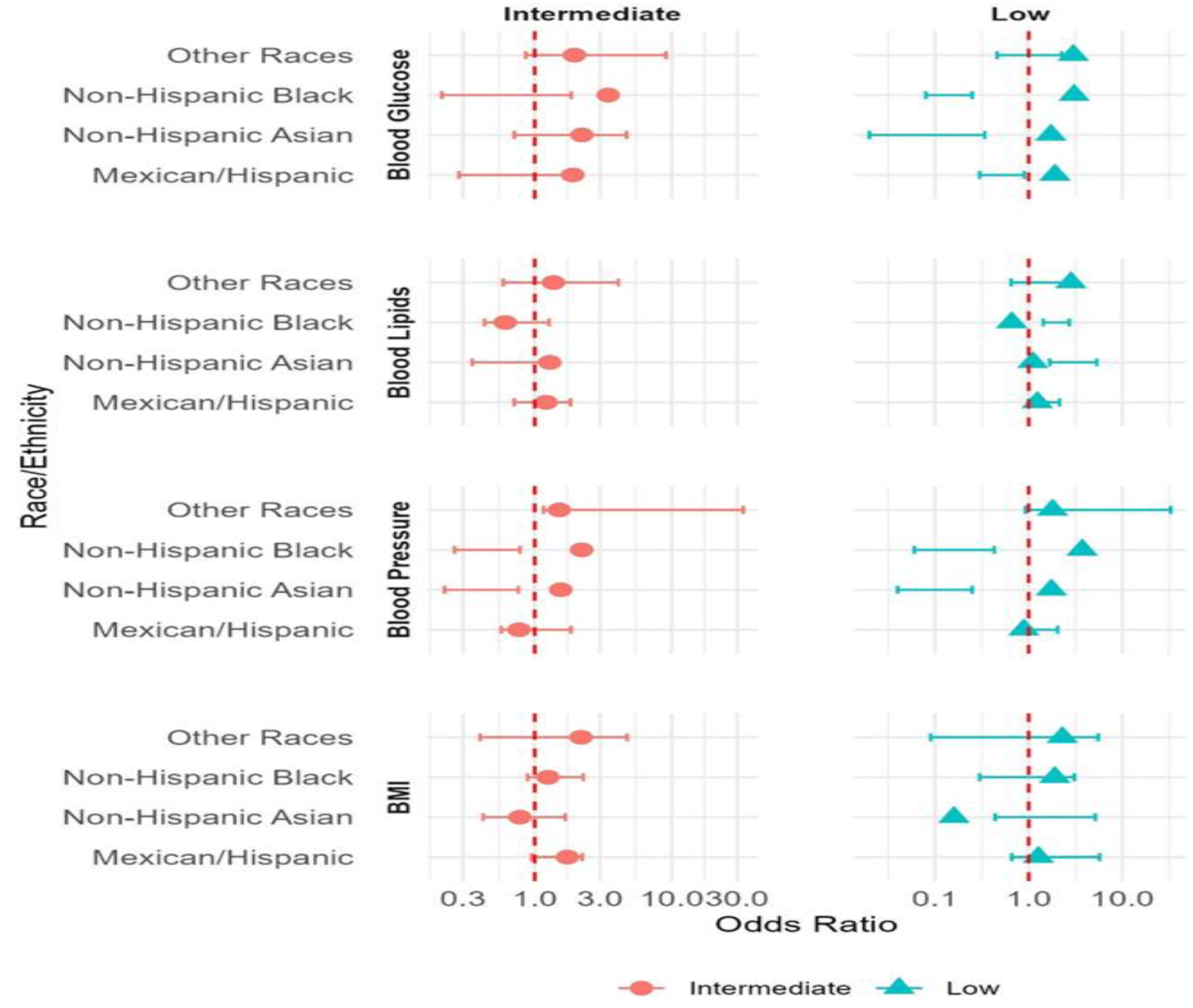
Adjusted Prevalence Rate Ratios of Life’s Essential 8 Health Factor Components by Race and Ethnicity Among Ever- Pregnant Adults (NHANES 2011-2020) Note: This figure displays adjusted prevalence rate ratios (PRRs) with 95% confidence intervals for the health factor components of the Life’s Essential 8 (LE8) score by race and ethnicity among ever-pregnant adults. The components shown are blood pressure (BP), blood glucose, blood lipids, and body mass index (BMI). Estimates are derived from survey-weighted multinomial logistic regression analyses using NHANES 2011-2020 data. The reference group is non-Hispanic White adults, and the reference category for each component is the “Ideal” level. PRRs above 1 indicate a higher likelihood, while PRRs below 1 indicate a lower likelihood of being in the “Intermediate” or “Poor” category than non- Hispanic Whites. Models are adjusted for age, education, poverty-income ratio, health insurance status, and employment. Whiskers represent 95% confidence intervals.

### Factors associated with Life’s Essential 8 score Quartiles among ever-pregnant adults

Factors associated with LE8 scores and LE8 quartiles are shown in **Table 4 and Table S4**. Participants in the older age categories were progressively less likely to be in higher LE8 score quartiles. Higher education was associated with higher LE8 score quartiles; those with ≥college education had 9.59 times higher odds (OR: 9.59, 95% CI: 6.16-14.92) of being in Quartile 4 compared to those with a high school education. Individuals with a PIR ≥2 had 1.10 times higher odds of being in the highest LE8 score quartile (OR: 1.10, 95% CI: 0.66-1.84) than those with a PIR <1. There were no significant associations between health insurance status or employment and LE8 score quartiles.

**Table 4:** Factors Associated with Life’s Essential 8 Score Quartiles Among Ever-Pregnant Adults (NHANES 2011–2020)

| **Variables** | **Quartile 1**<br>**(LE8 score: 29.28-53)** | **Quartile 2**<br>**(LE8 score: 53- 56.88)** | **Quartile 3**<br>**(LE8 score: 56.88 – 72.5)** | **Quartile 4**<br>**(LE8 score** |
| --- | --- | --- | --- | --- |
| **Race/ethnicity** |  |  |  |  |
| Non-Hispanic White [ref] | 1.00 | 1[1.00,1.00] | 1[1.00,1.00] | 1[1.00,1.00] |
| Non-Hispanic Black | 1.00 | 0.95[0.53,1.69] | 0.56***[0.42,0.76] | 0.27***[0.16,0.45] |
| Mexican/Hispanic | 1.00 | 1.80*[1.01,3.22] | 1.32[0.92,1.91] | 1.46[0.85,2.51] |
| Non-Hispanic Asian | 1.00 | 1.74[0.76,3.99] | 2.41**[1.43,4.08] | 1.34[0.64,2.82] |
| Other Races | 1.00 | 1.35[0.42,4.40] | 0.53[0.18,1.55] | 0.29**[0.13,0.66] |
| **Age** |  |  |  |  |
| 20-29 [ref] | 1.00 | 1[1.00,1.00] | 1[1.00,1.00] | 1[1.00,1.00] |
| 30-39 | 1.00 | 0.62[0.30,1.31] | 0.28***[0.15,0.50] | 0.42*[0.19,0.92] |
| 40-49 | 1.00 | 0.42*[0.21,0.81] | 0.27***[0.15,0.47] | 0.25**[0.11,0.58] |
| 50-59 | 1.00 | 0.41*[0.18,0.94] | 0.12***[0.06,0.27] | 0.14***[0.06,0.34] |
| 60-69 | 1.00 | 0.39*[0.17,0.86] | 0.17***[0.10,0.29] | 0.11***[0.06,0.20] |
| 70-79 | 1.00 | 0.56[0.23,1.34] | 0.16***[0.11,0.25] | 0.05***[0.02,0.13] |
| **Education** |  |  |  |  |
| High School [ref] | 1.00 | 1[1.00,1.00] | 1[1.00,1.00] | 1[1.00,1.00] |
| Some College | 1.00 | 1.441[0.89,2.34] | 1.58**[1.16,2.15] | 2.65***[1.57,4.48] |
| College graduate and above | 1.00 | 1.65[0.87,3.12] | 2.41***[1.71,3.40] | 9.59**[6.16,14.92] |
| **Poverty Index Ratio** |  |  |  |  |
| PIR <1 [ref] | 1.00 | 1[1.00,1.00] | 1[1.00,1.00] | 1[1.00,1.00] |
| PIR 1-1.99 | 1.00 | 0.87[0.54,1.40] | 1.05[0.77,1.43] | 0.71[0.45,1.11] |
| PIR ≥2 | 1.00 | 0.96[0.61,1.50] | 1.54*[1.10,2.16] | 1.10[0.66,1.84] |
| **Insurance** |  |  |  |  |
| No Health Insurance [ref] | 1.00 | 1[1.00,1.00] | 1[1.00,1.00] | 1[1.00,1.00] |
| Health Insurance | 1.00 | 1.16[0.65,2.07] | 0.97[0.73,1.27] | 0.99[0.54,1.82] |
| **Employment** |  |  |  |  |
| Unemployed [ref] | 1.00 | 1[1.00,1.00] | 1[1.00,1.00] | 1[1.00,1.00] |
| Employed | 1.00 | 1.20[0.73,1.96] | 1.25[0.82,1.88] | 0.99[0.58,1.69] |

### Sensitivity Analysis

In sensitivity analyses, participants of ‘other’ races were excluded; the fully adjusted multinomial logistic regression models showed consistent results (**Table S5**).

## DISCUSSION

In this nationally representative sample of ever-pregnant individuals, our findings show disparities in LE8 scores across groups, emphasizing the influence of race and ethnicity (as a proxy for racial inequities), age, and socioeconomic factors. Our results showed notable differences in overall LE8 scores and individual cardiovascular health components across racial and ethnic groups, with NH Black adults generally showing less favorable cardiovascular health profiles. We also found that older age, lower educational attainment, and lower income were associated with lower LE8 scores. Additionally, our analysis demonstrated significant variations in the prevalence of ideal levels for specific health behaviors and factors, such as blood pressure, blood glucose, HbA1c, and physical activity, across different racial and ethnic groups. These disparities in cardiovascular health underscore the urgent need for targeted interventions and policy changes to address the root causes of health inequities.

We found that NH Black ever-pregnant adults consistently showed lower LE8 scores, even in the lowest quartile, indicating a higher burden of cardiovascular risk factors in this population compared to NH Whites. These findings align with previous studies that have consistently reported higher rates of hypertension, obesity, diabetes, and other cardiometabolic conditions among NH Black populations compared to their NH White adults.^17–20^ This persistent disparity highlights the need for a more nuanced understanding of cardiovascular health beyond individual risk factors and encompasses the broader social and environmental determinants of health^21^. A complex interplay of individual, community, and societal factors may contribute to health disparities. Moreover, the intersectionality of race, gender, and pregnancy history in our study population adds another layer of complexity to these disparities, suggesting that interventions must be tailored to address the unique needs of women most impacted across their reproductive lifespan.^22^

The observed disparities in LE8 scores reflect differences in health outcomes and disparities in access to resources that promote cardiovascular health. For instance, lower scores in physical activity and diet quality among certain racial/ethnic groups may indicate limited access to safe outdoor spaces for exercise or healthy food options in underserved communities^23^. Similarly, disparities in sleep health may be linked to occupational factors, neighborhood characteristics, or chronic stress associated with experiences of discrimination^24^. Addressing these upstream factors is crucial for reducing cardiovascular health inequities. Addressing the intersectionality of common risk factors, like hypertension and poverty, may also be essential in reducing the excess burden of cardiovascular disease morbidity in this population. Further, our findings underscore the importance of adopting a life course approach to cardiovascular health, recognizing that disparities observed in adulthood may have roots in early life experiences and exposures.

Disparities in hypertension were evident among ever-pregnant adults, with NH Black individuals consistently exhibiting a significantly higher likelihood, aligning with prior research.^21–24^ This disparity is particularly alarming given the critical role of blood pressure control in cardiovascular health, especially for women who have experienced pregnancy. Hypertensive disorders of pregnancy are known risk factors for future cardiovascular disease, and these findings suggest that NH Black women may face a double burden: increased risk during pregnancy and decreased likelihood of optimal blood pressure control afterward.

Similarly, disparities in high blood glucose among NH Black and Mexican/Hispanic individuals aligned with earlier studies. ^25,26^ The persistent nature of this disparity suggests ongoing systemic issues in healthcare delivery and access for NH Black individuals, including discriminatory delivery of high-quality care. Disparities in lifestyle factors, including poor diet, inadequate physical activity, poor sleep, and overweight/obesity, were also identified. These findings align with previous studies highlighting the enduring challenges in promoting healthy behaviors.

Our study highlights the need for innovative approaches to address cardiovascular health disparities. Digital health interventions, including mobile health applications and wearable devices, offer promising avenues for promoting cardiovascular health across diverse populations.^31^ These technologies can provide personalized health information, track progress, and offer real-time support, potentially overcoming some barriers to healthcare access and health education^32^. However, ensuring that digital solutions are designed and implemented in ways that do not exacerbate existing disparities is crucial^33^. Future research should explore how to effectively leverage these technologies to improve cardiovascular health outcomes, particularly among populations most affected by disparities.

Additionally, our findings highlight the importance of addressing cardiovascular health within the broader context of reproductive health. The period before, during, and after pregnancy represents a critical window of opportunity for cardiovascular health promotion.^34^ Integrating cardiovascular health assessments and interventions into routine prenatal and postpartum care could have far-reaching impacts on women’s long-term health outcomes.^35^ Indeed, there is growing recognition of how important preconception health care is in shaping maternal and child health outcomes^36^. Overall, these findings advocate for using the LE8 score as a screening tool to identify high-risk women and implement multi-pronged interventions that address both individual behaviors and social determinants of health. By doing so, we can perhaps work towards reducing the burden of cardiometabolic complications and improving maternal health outcomes for all women.

### Limitations

Our study had several limitations that should be considered when interpreting the results. First, the inclusion criterion of a broad spectrum of women’s life stages extending from young adulthood to women in their 60s and beyond presents a limitation. The wide age range may not adequately differentiate the specific cardiovascular health concerns of women of reproductive age from those of older, postmenopausal women. Second, many LE8 components use self-reported data, which may be subject to recall or social desirability bias. Our study focused on ever- pregnant adults, which may limit the applicability of our findings to nulliparous women or men. Additionally, our study did not exclusively focus on women of reproductive age, which could introduce variability in cardiovascular risk factors across different life stages. We also did not account for the time elapsed since pregnancy, which may impact the observed associations. Lastly, while the LE8 score provides a comprehensive measure of cardiovascular health, it may not capture all relevant aspects of cardiovascular risk, particularly those specific to women’s reproductive health. Future research should consider incorporating additional metrics that may be particularly relevant to women’s cardiovascular health across the lifespan.

## Conclusions

Our study shows significant disparities in cardiovascular health among ever-pregnant adults in the United States, with particularly concerning trends for NH Black women. These findings also show the complex interplay of individual, social, and structural factors shaping cardiovascular health across the life course. Addressing these disparities requires a multifaceted approach, integrating comprehensive cardiovascular care into women’s health services, leveraging technological innovations, and implementing community-based interventions. Importantly, we must focus on equity and cultural competence in all initiatives and work towards reducing the burden of cardiovascular disease and improving health outcomes for the most impacted populations. Our findings call for urgent action to create a healthier, more equitable future for all women and their families.

## Non-standard Abbreviations and Acronyms

LE8: Life’s Essential 8
NHANES: National Health and Nutrition Examination Survey
NH: Non-Hispanic
PRR: Prevalence Rate Ratio
PIR: Poverty Income Ratio
AHA: American Heart Association
BMI: Body Mass Index
HbA1c: Hemoglobin A1c

## Data Availability

Data is publicly available.

## ACKNOWLEDGMENTS

We thank the National Health and Nutrition Examination Survey participants, staff, and the National Center for Health Statistics for their valuable contributions.

## Prior Presentation

The abstract was presented at the 2023 American Heart Association Hypertension Meeting.

## SUPPLEMENTAL MATERIALS

**Figure S1:**
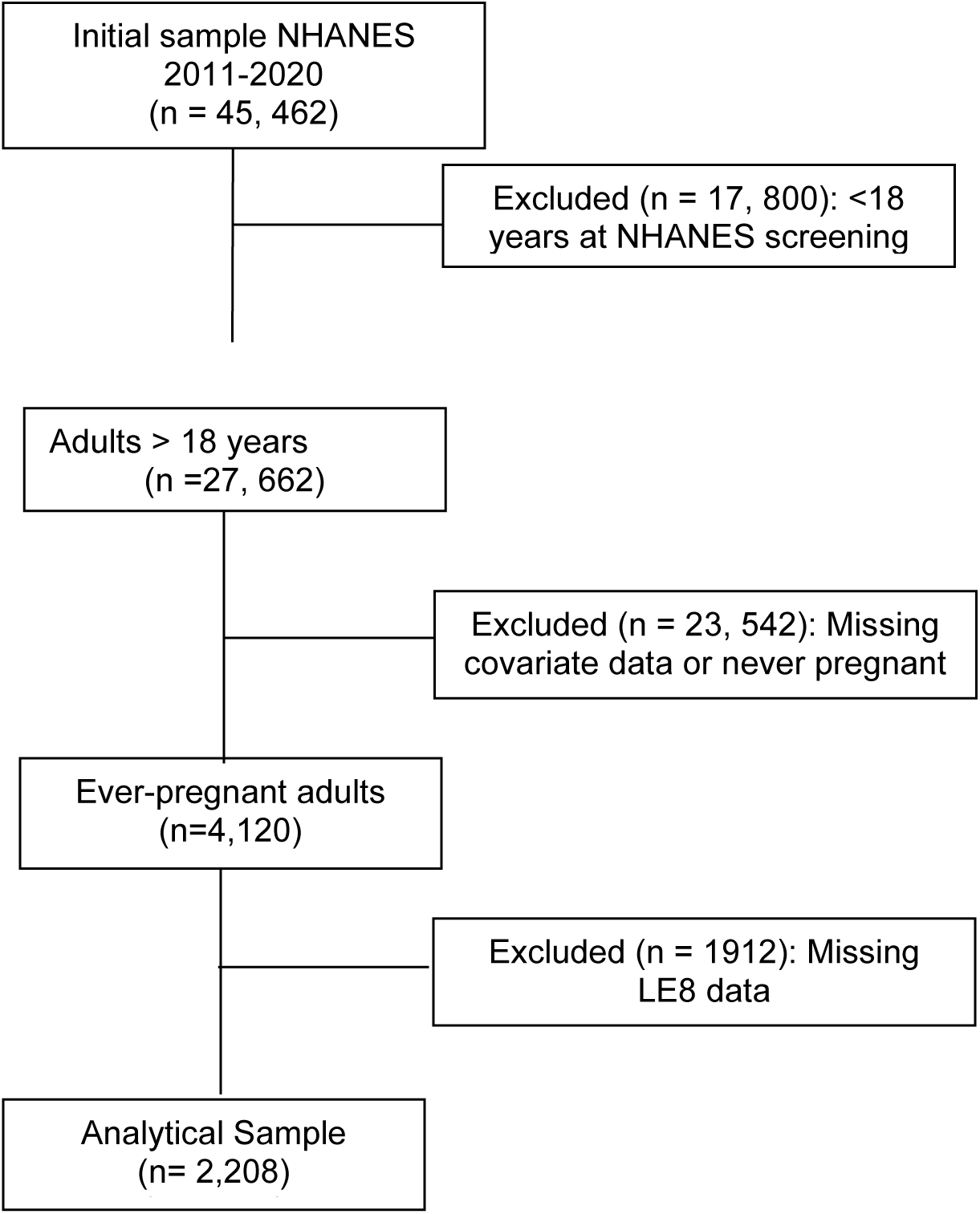
Flow Diagram for Study Participant Selection.

**Table S1:**
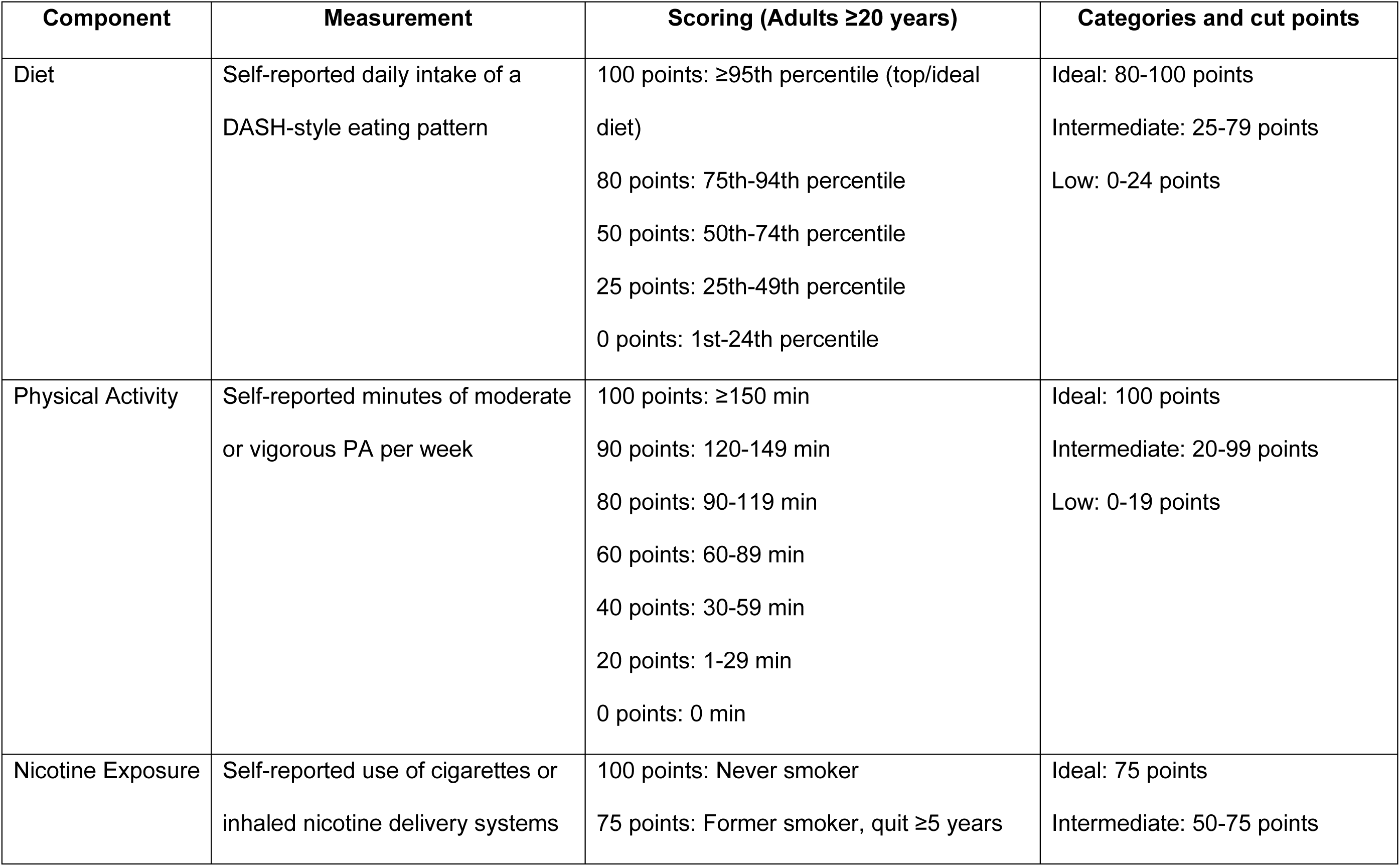
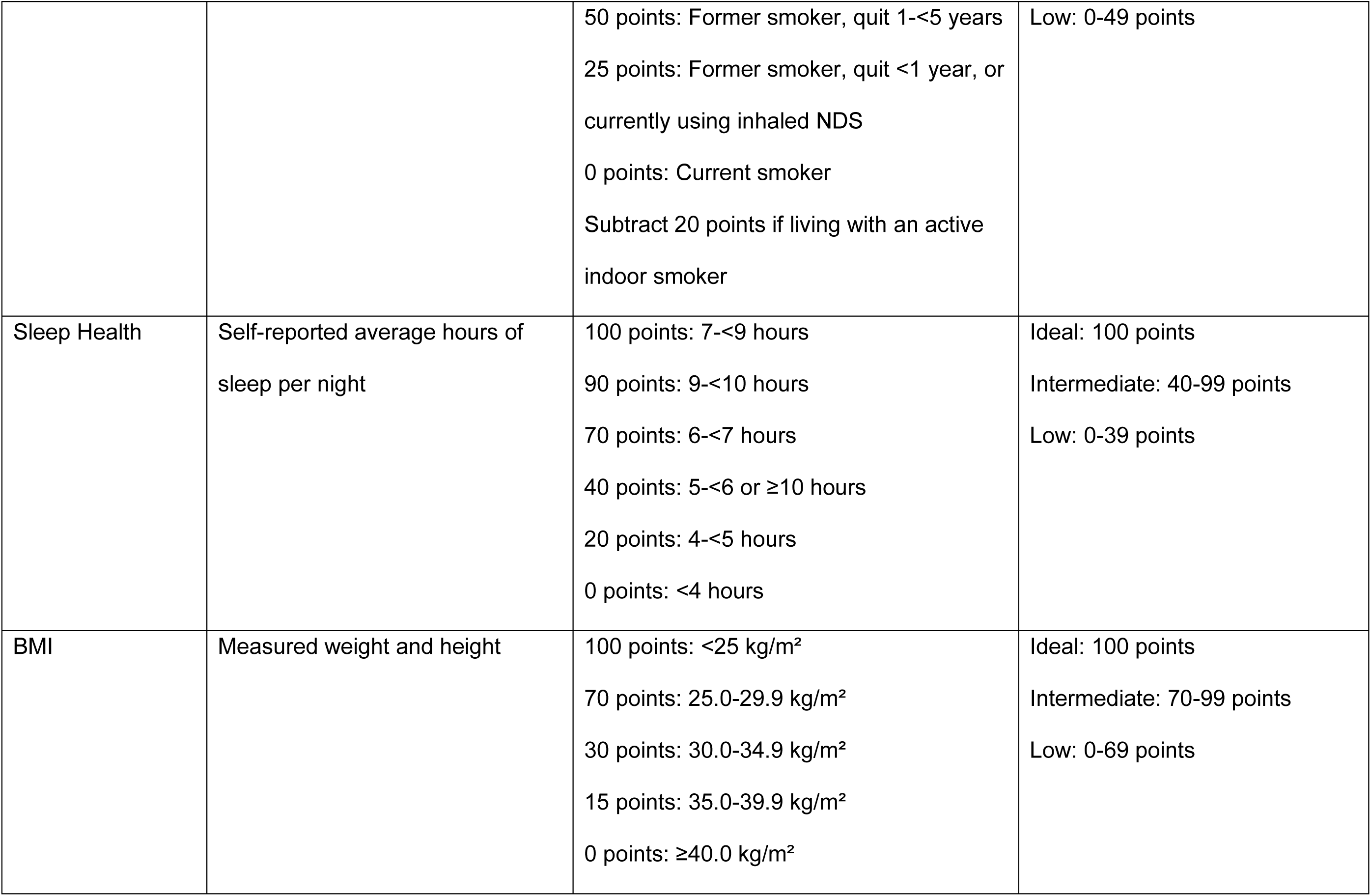
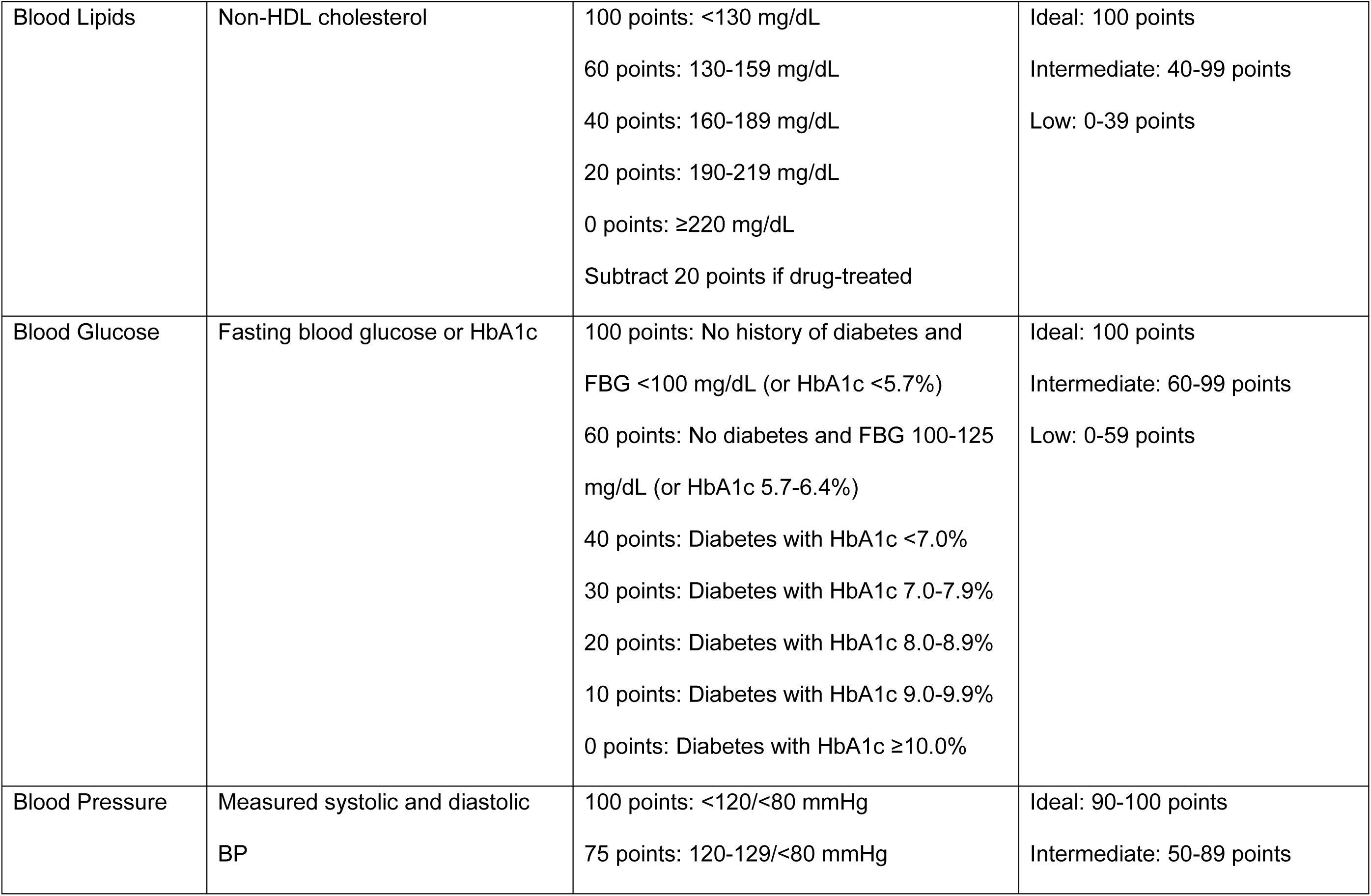
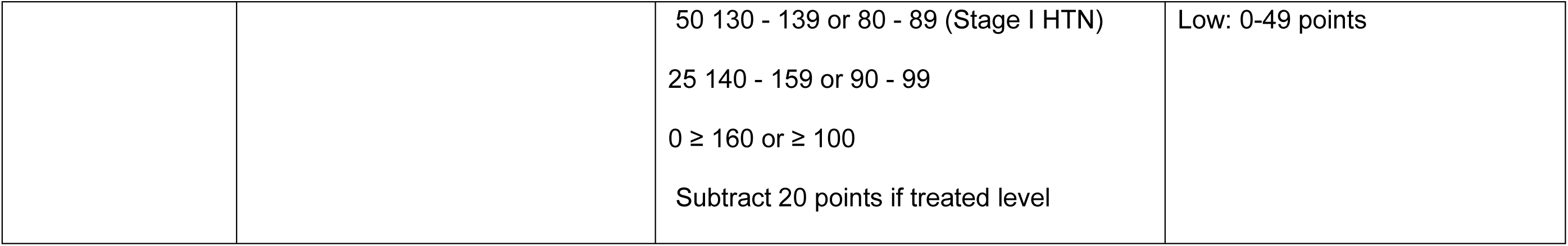
Life’s Essential 8 Score Calculations Using NHANES 2011-2020 Data.

| **Component** | **Measurement** | **Scoring (Adults ≥20 years)** | **Categories and cut points** |
| --- | --- | --- | --- |
| Diet | Self-reported daily intake of a DASH-style eating pattern | 100 points: ≥95th percentile (top/ideal diet)<br>80 points: 75th-94th percentile<br>50 points: 50th-74th percentile<br>25 points: 25th-49th percentile<br>0 points: 1st-24th percentile | Ideal: 80-100 points<br>Intermediate: 25-79 points<br>Low: 0-24 points |
| Physical Activity | Self-reported minutes of moderate or vigorous PA per week | 100 points: ≥150 min<br>90 points: 120-149 min<br>80 points: 90-119 min<br>60 points: 60-89 min<br>40 points: 30-59 min<br>20 points: 1-29 min<br>0 points: 0 min | Ideal: 100 points<br>Intermediate: 20-99 points<br>Low: 0-19 points |
| Nicotine Exposure | Self-reported use of cigarettes or inhaled nicotine delivery systems | 100 points: Never smoker<br>75 points: Former smoker, quit ≥5 years | Ideal: 75 points<br>Intermediate: 50-75 points |
|  |  | 50 points: Former smoker, quit 1-<5 years<br>25 points: Former smoker, quit <1 year, or currently using inhaled NDS<br>0 points: Current smoker<br>Subtract 20 points if living with an active indoor smoker | Low: 0-49 points |
| Sleep Health | Self-reported average hours of sleep per night | 100 points: 7-<9 hours<br>90 points: 9-<10 hours<br>70 points: 6-<7 hours<br>40 points: 5-<6 or ≥10 hours<br>20 points: 4-<5 hours<br>0 points: <4 hours | Ideal: 100 points<br>Intermediate: 40-99 points<br>Low: 0-39 points |
| BMI | Measured weight and height | 100 points: <25 kg/m<sup>2</sup><br>70 points: 25.0-29.9 kg/m<sup>2</sup><br>30 points: 30.0-34.9 kg/m<sup>2</sup><br>15 points: 35.0-39.9 kg/m<sup>2</sup><br>0 points: ≥40.0 kg/m<sup>2</sup> | Ideal: 100 points<br>Intermediate: 70-99 points<br>Low: 0-69 points |
| Blood Lipids | Non-HDL cholesterol | 100 points: <130 mg/dL<br>60 points: 130-159 mg/dL<br>40 points: 160-189 mg/dL<br>20 points: 190-219 mg/dL<br>0 points: ≥220 mg/dL<br>Subtract 20 points if drug-treated | Ideal: 100 points<br>Intermediate: 40-99 points<br>Low: 0-39 points |
| Blood Glucose | Fasting blood glucose or HbA1c | 100 points: No history of diabetes and<br>FBG <100 mg/dL (or HbA1c <5.7%)<br>60 points: No diabetes and FBG 100-125<br>mg/dL (or HbA1c 5.7-6.4%)<br>40 points: Diabetes with HbA1c <7.0%<br>30 points: Diabetes with HbA1c 7.0-7.9%<br>20 points: Diabetes with HbA1c 8.0-8.9%<br>10 points: Diabetes with HbA1c 9.0-9.9%<br>0 points: Diabetes with HbA1c ≥10.0% | Ideal: 100 points<br>Intermediate: 60-99 points<br>Low: 0-59 points |
| Blood Pressure | Measured systolic and diastolic<br>BP | 100 points: <120/<80 mmHg<br>75 points: 120-129/<80 mmHg | Ideal: 90-100 points<br>Intermediate: 50-89 points |
| | | 50 130 - 139 or 80 - 89 (Stage I HTN)<br>25 140 - 159 or 90 - 99<br>0 ≥ 160 or ≥ 100<br>Subtract 20 points if treated level | Low: 0-49 points |

**Table S2:** Unweighted Sample Distribution of Ever pregnant Adults by Race and Ethnicity (N=2,208)

|  |  |  |  |  |  |  |
| --- | --- | --- | --- | --- | --- | --- |
| **Variable n (%)** | **Non-Hispanic**<br>**White** | **Mexican**<br>**American/Other**<br>**Hispanic** | **Non-Hispanic Black** | **Non-Hispanic Asian** | **Other Races** | **Total** |
| **N** | 844 (38.23) | 482 (21.83) | 606 (27.45) | 184 (8.33) | 92 (4.17) | 2208 (100) |
| Age, yr (mean, SD) | 54.43(±14.70) | 45.42(±22.64) | 48.03(±30.05) | 49.79(±25.10) | 48.12(±18.96) | 52.00(±19.64) |
| **Age Categories -10yr intervals** |  |  |  |  |  |  |
| 20-29 years | 52 (6.16) | 57 (11.83) | 74 (12.21) | 10 (5.43) | 14 (15.21) | 207 (9.38) |
| 30-39 years | 131 (15.52) | 91 (18.88) | 99 (16.34) | 35 (19.02) | 16 (17.39) | 372 (16.85) |
| 40-49 years | 132 (15.64) | 93 (19.30) | 109 (17.99) | 42 (22.83) | 23 (25) | 399 (18.07) |
| 50-59 years | 143 (16.94) | 104 (21.58) | 119 (19.64) | 51 (27.72) | 15 (16.30) | 432 (19.57) |
| 60-69 years | 157 (18.60) | 91 (18.88) | 144 (23.76) | 29 (15.76) | 17 (18.48) | 438 (19.84) |
| 70+ years | 229 (27.13) | 46 (9.54) | 61 (10.07) | 17 (9.24) | 7 (7.61) | 360 (16.30) |
| **Education** |  |  |  |  |  |  |
| High School | 288 (34.12) | 275 (57.05) | 229 (37.79) | 38 (20.65) | 27 (29.35) | 857 (38.81) |
| Some College | 353 (41.83) | 145 (30.08) | 250 (41.25) | 42 (22.83) | 50 (54.35) | 840 (38.04) |
| College graduate<br>and above | 203 (24.05) | 62 (12.86) | 127 (20.96) | 104 (56.52) | 15 (16.30) | 511 (23.14) |
| **Poverty-Income Ratio** |  |  |  |  |  |  |
| PIR <1 | 112 (13.27) | 130 (26.97) | 177 (29.21) | 9 (4.89) | 25 (27.17) | 453 (20.52) |
| PIR 1-1.99 | 220 (26.07) | 152 (31.54) | 177 (29.21) | 28 (15.22) | 28 (30.44) | 605 (27.40) |
| PIR ≥2 | 512 (60.66) | 200 (41.49) | 252 (41.58) | 147 (79.89) | 39 (42.39) | 1150 (52.08) |
| **Health Insurance Status** |  |  |  |  |  |  |
| No Health<br>Insurance | 69 (8.18) | 114 (23.65) | 86 (14.19) | 16 (8.69) | 8 (8.69) | 293 (13.27) |
| Have Health<br>Insurance | 775 (91.83) | 368 (76.35) | 520 (85.81) | 168 (91.30) | 84 (91.30) | 1915 (86.73) |
| **Employment Status** |  |  |  |  |  |  |
| Unemployed | 477 (56.52) | 246 (51.04) | 277 (45.71) | 73 (39.67) | 43 (46.74) | 1116 (50.54) |
| Employed | 367 (43.48) | 236 (48.96) | 329 (54.29) | 111 (60.33) | 49 (53.26) | 1092 (49.46) |
| All significant at P<0.001 |  |  |  |  |  |  |

**Table S3:** Racial/Ethnic Differences in Life’s Essential 8 Components Among Ever-Pregnant Adults, NHANES 2011-2020.

| LE8 Component | Race and Ethnicity Categories |  |  |  |  |
| --- | --- | --- | --- | --- | --- |
|  | Non-Hispanic White | Non-Hispanic Black | Mexican/Hispanic | Non-Hispanic Asian | Other Races |
| **Overall LE8 Score** |  |  |  |  |  |
| Moderate | 1.00 (Ref) | 0.61** (0.44, 0.87) | 1.56 (0.97, 2.50) | 2.18** (1.23, 3.89) | 0.73 (0.36, 1.45) |
| High | 1.00 (Ref) | 0.20*** (0.10, 0.42) | 1.02 (0.53, 1.99) | 1.18 (0.53, 2.63) | 0.23* (0.08, 0.71) |
| **Blood Pressure** |  |  |  |  |  |
| Intermediate | 1.00 (Ref) | 2.20***[1.67,2.89] | 0.77[0.52,1.13] | 1.55*[1.02,2.36] | 1.51[0.62,3.71] |
| Low | 1.00 (Ref) | 3.73***[2.56,5.42] | 0.89[0.63,1.26] | 1.75*[1.04,2.96] | 1.80[0.75,4.29] |
| **Blood Glucose** |  |  |  |  |  |
| Intermediate | 1.00 (Ref) | 3.44***[2.85,4.14] | 1.90***[1.42,2.55] | 2.21**[1.39,3.52] | 1.95*[1.03,3.69] |
| Low | 1.00 (Ref) | 3.06***[2.05,4.57] | 1.91**[1.19,3.05] | 1.73 (0.84, 3.58) | 3.00[0.99,9.12] |
| **Blood Lipids** |  |  |  |  |  |
| Intermediate | 1.00 (Ref) | 0.61*[0.41,0.89] | 1.21[0.94,1.56] | 1.29[0.88,1.88] | 1.37[0.71,2.65] |
| Low | 1.00 (Ref) | 0.66*[0.46,0.95] | 1.24[0.70,2.18] | 1.11[0.58,2.11] | 2.83[0.94,8.55] |
| **BMI** |  |  |  |  |  |
| Intermediate | 1.00 (Ref) | 1.25[0.91,1.72] | 1.73**[1.25,2.38] | 0.78[0.42,1.45] | 2.18[0.89,5.33] |
| Low | 1.00 (Ref) | 1.90***[1.44,2.52] | 1.27[0.94,1.72] | 0.16***[0.09,0.28] | 2.29**[1.25,4.22] |
| **Sleep** |  |  |  |  |  |
| Intermediate | 1.00 (Ref) | 1.37[0.94,1.99] | 1.48[0.93,2.33] | 0.91[0.47,1.76] | 1.28[0.42,3.95] |
| Low | 1.00 (Ref) | 1.55[0.54,4.48] | 0.92[0.29,2.93] | 1.31[0.38,4.55] | 0.46[0.06,3.55] |
| **Smoking** |  |  |  |  |  |
| Intermediate | 1.00 (Ref) | 0.47[0.20,1.09] | 0.51[0.23,1.13] | 1.06[0.38,2.96] | 2.21*[1.02,4.79] |
| Low | 1.00 (Ref) | 0.54*[0.32,0.92] | 0.14***[0.08,0.26] | 0.08**[0.02,0.32] | 1.04[0.49,2.18] |
| **Physical Activity** |  |  |  |  |  |
| Intermediate | 1.00 (Ref) | 1.16[0.73,1.85] | 0.47**[0.27,0.81] | 0.79[0.42,1.47] | 1.49[0.58,3.84] |
| Low | 1.00 (Ref) | 1.40[0.97,2.03] | 1.75**[1.26,2.41] | 2.85***[1.65,4.91] | 1.37[0.76,2.46] |
| **Diet** |  |  |  |  |  |
| Intermediate | 1.00 (Ref) | 1.09[0.62,1.91] | 0.45** (0.26, 0.78) | 0.48*[0.26,0.88] | 6.55*[1.22,35.35] |
| Low | 1.00 (Ref) | 1.27[0.80,2.03] | 0.15***[0.06,0.38] | 0.12***[0.05,0.31] | 5.96*[1.03,34.64] |
| Values are presented as prevalence rate ratios with 95% confidence intervals in parentheses. The reference category for each component is the “Ideal” category. The reference group for race/ethnicity is Non-Hispanic White.<br>p<0.05, ** p<0.01, *** p<0.001 |  |  |  |  |  |

**Table S4:** Factors associated with Life’s Essential 8 scores among ever-pregnant adults (NHANES 2011–2020)

|  |  |
| --- | --- |
| **Race/Ethnicity** | **LE8 Score** |
| Non-Hispanic White[ref] | 1.00 |
| Non-Hispanic Black | 0.01***[0.00,0.02] |
| Mexican/Hispanic | 3.25[0.41,25.90] |
| Non-Hispanic Asian | 2.55[0.27,24.23] |
| Other Races | 0.01*[0.00,0.34] |
| **Age** |  |
| 20-29[ref] | 1.00 |
| 30-49 | 0.03**[0.00,0.40] |
| 40-49 | 0.01**[0.00,0.13] |
| 50-69 | 0.00***[0.00,0.00] |
| 60-69 | 0.00***[0.00,0.00] |
| 70-79 | 0.00***[0.00,0.00] |
| **Education** |  |
| High School[ref] | 1.00 |
| Some College | 16.61**[2.96,93.36] |
| College graduate and above | 5576.1***[1144.00,27178.97] |
| **Poverty Index Ratio** |  |
| PIR <1 [ref] | 1.00 |
| PIR 1 - 1.9 | 0.74[0.16,3.31] |
| PIR ≥2 | 9.31*[1.26,68.54] |
| **Insurance** |  |
| No Health insurance[ref] | 1.00 |
| Health insurance | 0.51[0.06,4.17] |
| **Employment**[ref] |  |
| Unemployed | 1.00 |
| Employed | 0.99[0.13,7.68] |
| Values are presented as beta coefficients from linear regression with 95% confidence intervals. * p<0.05, ** p<0.01, *** p<0.001 |  |

**Table S5:** Sensitivity Analysis - Factors Associated with Life’s Essential 8 Score Quartiles Among Ever-Pregnant Adults (Excluding “Other Races”), NHANES 2011-2020.

|  | **Quartile 1**<br>(LE8 score: 25-47.5) | **Quartile 2**<br>(LE8 score: 47.5- 51.14) | **Quartile 3**<br>(LE8 score: 51.14- 67.5) | **Quartile 4**<br>(LE8 score: ≥67.5) |
| --- | --- | --- | --- | --- |
| **Race** |  |  |  |  |
| Non-Hispanic White<br>[ref] | 1.00 | 1.00 | 1.00 | 1.00 |
| Non-Hispanic Black | 1.00 | 0.95[0.54,1.67] | 0.57***[0.43,0.77] | 0.28***[0.17,0.45] |
| Mexican/Hispanic | 1.00 | 1.86*[1.05,3.29] | 1.34[0.94,1.92] | 1.49[0.88,2.52] |
| Non-Hispanic Asian | 1.00 | 1.79[0.78,4.14] | 2.41**[1.43,4.08] | 1.33[0.63,2.82] |
| **Age** |  |  |  |  |
| 20-29 [ref] | 1.00 | 1.00 | 1.00 | 1.00 |
| 30-39 | 1.00 | 0.61[0.29,1.30] | 0.28***[0.14,0.53] | 0.44[0.19,1.04] |
| 40-49 | 1.00 | 0.35**[0.16,0.76] | 0.30***[0.16,0.54] | 0.27**[0.11,0.65] |
| 50-59 | 1.00 | 0.36*[0.16,0.80] | 0.13***[0.05,0.29] | 0.14***[0.06,0.36] |
| 60-69 | 1.00 | 0.40*[0.18,0.89] | 0.17***[0.09,0.31] | 0.11***[0.05,0.22] |
| 70-79 | 1.00 | 0.52[0.20,1.35] | 0.17***[0.11,0.27] | 0.05***[0.02,0.14] |
| **Education** |  |  |  |  |
| High School [ref] | 1.00 | 1.00 | 1.00 | 1.00 |
| Some College | 1.00 | 1.65[0.96,2.84] | 1.56**[1.15,2.13] | 2.61***[1.55,4.38] |
| College graduate and above | 1.00 | 1.66[0.83,3.31] | 2.37***[1.71,3.26] | 9.37***[6.01,14.59] |
| **Poverty Index Ratio** |  |  |  |  |
| PIR <1 [ref] | 1.00 | 1.00 | 1.00 | 1.00 |
| PIR 1-1.99 | 1.00 | 0.84[0.50,1.42] | 1.18[0.86,1.61] | 0.77[0.48,1.21] |
| PIR ≥2 | 1.00 | 0.92[0.57,1.48] | 1.71**[1.21,2.42] | 1.21[0.72,2.03] |
| **Insurance** |  |  |  |  |
| No Health Insurance [ref] | 1.00 | 1.00 | 1.00 | 1.00 |
| Health Insurance | 1.00 | 1.35[0.72,2.52] | 0.97[0.74,1.27] | 1.02[0.54,1.91] |
| **Employment** |  |  |  |  |
| Unemployed [ref] | 1.00 | 1.00 | 1.00 | 1.00 |
| Employed | 1.00 | 1.27[0.76,2.11] | 1.24[0.82,1.87] | 0.97[0.57,1.65] |
| Values are presented as odds ratios from multinomial logistic regression with 95% confidence intervals. The reference category for LE8 score quartiles is Quartile 1 (lowest scores). * p<0.05, ** p<0.01, *** p<0.001 |  |  |  |  |

